# Did financial interventions offset the impact of financial adversity on mental health during the COVID-19 pandemic? A longitudinal analysis of the UCL COVID-19 Social Study

**DOI:** 10.1101/2022.11.15.22282337

**Authors:** Elise Paul, Daisy Fancourt

## Abstract

**Background:** It remains unclear whether financial support interventions (e.g., furlough, mortgage freezes, foodbanks, Universal Credit) provide protection against the negative impact of financial adversity on mental health.

**Methods:** Data were from adults who took part in the UCL COVID -19 Social Study between 1 April 2020 and 4 April 2022 who had variability over time in depression (N = 27,297) and anxiety symptoms (N = 26,452). Fixed-effects Poisson regressions examined the associations between an index of financial adversity (e.g., job or income loss) with depression and anxiety symptoms and controlled for other adversities and loneliness. Interaction terms between financial adversity and having used i) any, ii) charity based, iii) government based, iv) work based, and v) other forms of financial supports were examined.

**Results:** Experiencing financial adversity had a negative impact on mental health. Only charity based support (e.g., foodbanks) consistently attenuated the impact of financial adversity on mental health, whilst work based support exacerbated the impact. Government based support only attenuated the impact of facing limited financial adversity on depression symptoms.

**Conclusion:** Findings suggest that most financial interventions are insufficient for alleviating mental health difficulties resulting from financial adversity.

## Introduction

Financial adversities such as loss of income or paid work have a negative impact on mental health;^1^ a finding that has been confirmed during the COVID-19 pandemic^2,3^. Identifying financial interventions and supports that may offset adverse psychological impacts is therefore necessary for allocating public resources. In the earlier stages of the pandemic, furlough was associated with declines in mental and social wellbeing in nine UK studies, but these declines were smaller than in people who were no longer in work.^4^ A US study found that amongst individuals experiencing financial adversity, living in a state with more generous social policies (e.g., medical coverage and unemployment benefits) buffered the impact on depression and anxiety.^5^ Before the pandemic, the negative impact of reducing government spending on social welfare (by consolidating multiple benefit schemes into Universal Credit) in the UK on mental health has also been documented.^6^ But in-depth analyses of different types of financial support schemes and their effects on mental health during COVID-19 are lacking.

We used data from a large, longitudinal dataset of measures repeated monthly to explore time-varying associations between the experience of financial adversity and anxiety and depression over a period of two years (April 2020 to April 2022). We then explored whether the impact of financial adversity on mental health was attenuated by the use of any financial support as well as four specific categories of financial support: charity, government, work based, and other.

## Methods

### Participants

We used data from the COVID-19 Social Study; a large panel study of the psychological and social experiences of over 70,000 adults (aged 18+) in the UK during the COVID-19 pandemic. The study commenced on 21 March 2020 and involved online weekly data collection through August 2020, and then monthly thereafter. Although not random, sampling utilised diverse methods resulting in a heterogeneous sample that was then weighted to UK population proportions. See Supplemental Materials for more information.

For these analyses, inclusion criteria were i) data on at least one of the four financial support modules (see Supplemental Table S1 for a listing of dates of administration), ii) data for at least two time points over the study period (1 April 2020 to 4 April 2022), iii) complete data on study variables, and iv) variability over time in depression and anxiety scores so that the associations between change in financial adversity and change in mental health could be analysed.

### Measures

#### Outcomes

Depression symptoms were measured monthly across the entirety of the study using the 9-item Patient Health Questionnaire (PHQ-9; range 0-27),^7^ and anxiety symptoms were assessed using the Generalized Anxiety Disorder Assessment (GAD-7; range 0-21).^8^ Both are standardised instruments used for depression and anxiety screening in primary care with higher scores indicating more symptoms.

#### Financial adversity

Financial adversity was operationalised as an index of five possible financial adversities (scored 0, 1, 2, 3, 4+): (i) loss of job/been unable to do paid work; (ii) spouse/partner lost their job/was unable to do paid work; (iii) major cut in household income (e.g., due to you or your partner being furloughed/put on leave/ not receiving sufficient work); (iv) unable to pay bills/rent/mortgage; and (v) evicted/lost accommodation. See Supplemental Table S3 for a full listing of items.

#### Financial supports

Modules enquiring about the use of various financial supports were administered a total of four times over the study period (Supplemental Table S1 for timing and stem questions and Supplemental Table S3 for response options and categorisations). Participants were invited to check all that applied from a list of 16 financial supports. These items were operationalised as five binary variables indicating having ever used any form of financial support, charity based support (e.g., foodbanks), government support (e.g., universal credit), work based support (e.g., furlough), and other supports (e.g., insurance claims) over the study period.

#### Time-varying covariates

Analyses controlled for (i) four other types of adversity: not being able to access essential items, loss of or serious illness in others, having been physically or psychologically abused, and infection with COVID-19 (see Supplemental Table S3); and (ii) loneliness was measured using the 3-item UCLA-3 Loneliness scale (range 0 hardly ever to 2 often).

## Statistical analysis

Fixed effects Poisson regression models were used to model associations between changes in financial adversity with changes in depression and anxiety symptoms over time whilst adjusting for time-varying covariates. These models were repeated with an interaction term between the financial adversity variable and the variables indicating financial support use. See Supplementary Materials for more detail, including model equation. Resulting regression coefficients were exponentiated and presented as incident rate ratios along with 95% confidence intervals.

To account for the non-random nature of the sample and increase representativeness of the UK general population, all data were weighted to the proportions of gender, age, ethnicity, country, and education obtained from the Office for National Statistics.^9^ Weights were constructed using a multivariate reweighting method using the Stata user written command ‘ebalance’.^10^ Analyses were conducted using Stata version 17.^11^

## Results

### Sample characteristics

Before sampling weights were applied, both samples were disproportionately female, of older age, and highly educated (Supplemental Table S2). However, after weights were applied, sample demographics better reflected those of the general UK population (Table 1). Nearly 40% of both samples had used at least one form of support at any point over the study period, with government support the most common type (22.27% and 22.53% in the depression and anxiety symptoms samples, respectively), followed by work based support (16.11% and 16.45% in the depression and anxiety symptoms samples, respectively). There was within-individual variation in both of the outcome variables and in the financial adversity measure (Table 2), confirming the appropriateness of using fixed effects models.

**Table 1.**
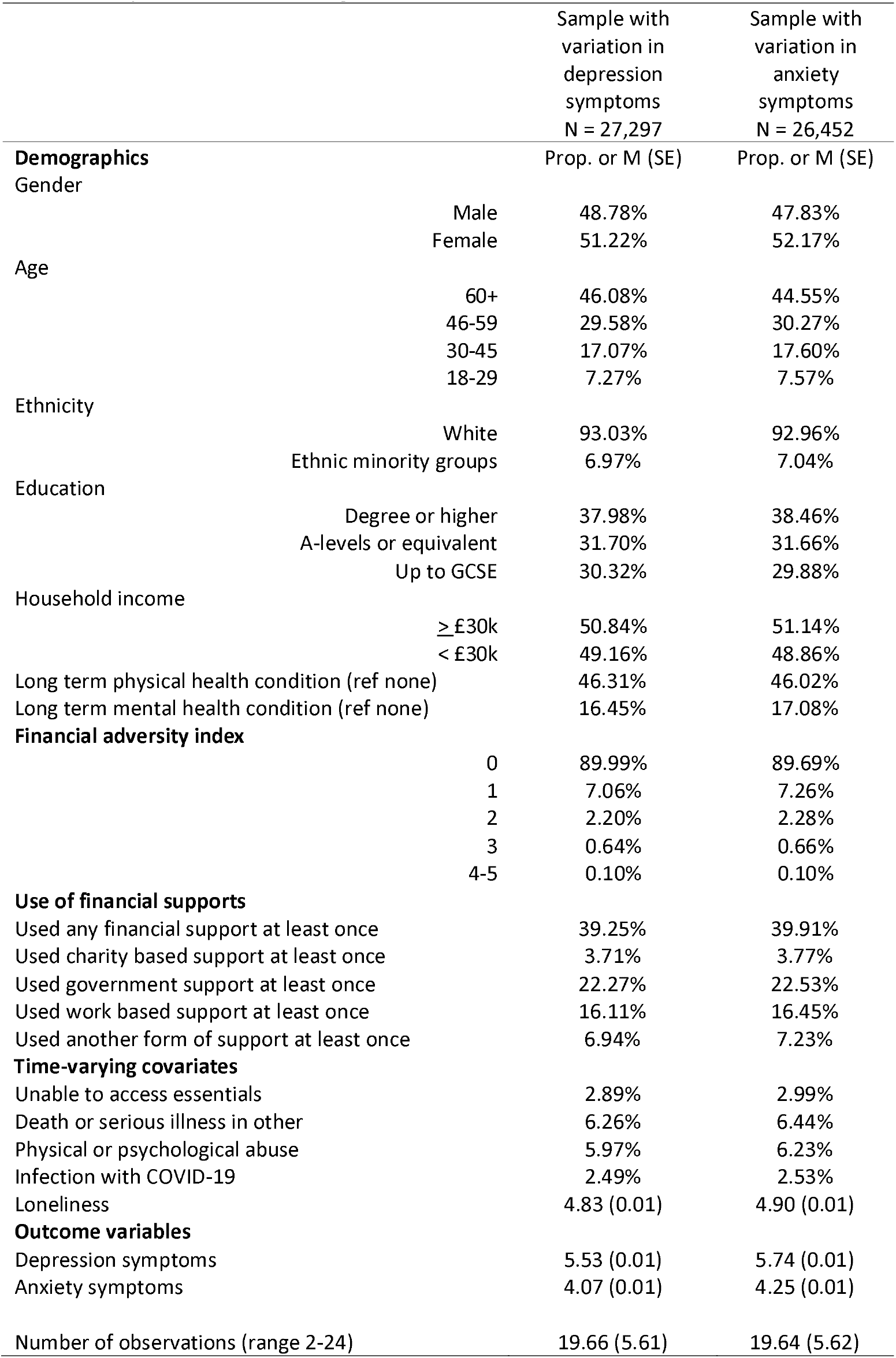
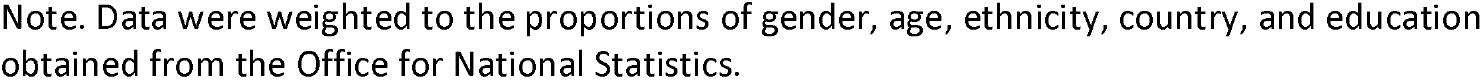
Sample characteristics, weighted ^a^

|  |  | Sample with<br>variation in<br>depression<br>symptoms<br>N = 27,297 | Sample with<br>variation in<br>anxiety<br>symptoms<br>N = 26,452 |
| --- | --- | --- | --- |
| **Demographics** |  | Prop. or M (SE) | Prop. or M (SE) |
| Gender | Male | 48.78% | 47.83% |
|  | Female | 51.22% | 52.17% |
| Age | 60+ | 46.08% | 44.55% |
|  | 46-59 | 29.58% | 30.27% |
|  | 30-45 | 17.07% | 17.60% |
|  | 18-29 | 7.27% | 7.57% |
| Ethnicity | White | 93.03% | 92.96% |
|  | Ethnic minority groups | 6.97% | 7.04% |
| Education | Degree or higher | 37.98% | 38.46% |
|  | A-levels or equivalent | 31.70% | 31.66% |
|  | Up to GCSE | 30.32% | 29.88% |
| Household income | ≥ £30k | 50.84% | 51.14% |
|  | < £30k | 49.16% | 48.86% |
| Long term physical health condition (ref none) |  | 46.31% | 46.02% |
| Long term mental health condition (ref none) |  | 16.45% | 17.08% |
| **Financial adversity index** |  |  |  |
| 0 |  | 89.99% | 89.69% |
| 1 |  | 7.06% | 7.26% |
| 2 |  | 2.20% | 2.28% |
| 3 |  | 0.64% | 0.66% |
| 4-5 |  | 0.10% | 0.10% |
| **Use of financial supports** |  |  |  |
| Used any financial support at least once |  | 39.25% | 39.91% |
| Used charity based support at least once |  | 3.71% | 3.77% |
| Used government support at least once |  | 22.27% | 22.53% |
| Used work based support at least once |  | 16.11% | 16.45% |
| Used another form of support at least once |  | 6.94% | 7.23% |
| **Time-varying covariates** |  |  |  |
| Unable to access essentials |  | 2.89% | 2.99% |
| Death or serious illness in other |  | 6.26% | 6.44% |
| Physical or psychological abuse |  | 5.97% | 6.23% |
| Infection with COVID-19 |  | 2.49% | 2.53% |
| Loneliness |  | 4.83 (0.01) | 4.90 (0.01) |
| **Outcome variables** |  |  |  |
| Depression symptoms |  | 5.53 (0.01) | 5.74 (0.01) |
| Anxiety symptoms |  | 4.07 (0.01) | 4.25 (0.01) |
| Number of observations (range 2-24) |  | 19.66 (5.61) | 19.64 (5.62) |

Note. Data were weighted to the proportions of gender, age, ethnicity, country, and education obtained from the Office for National Statistics.

**Table 2.** Within and between variation in the financial adversity index and outcome measures

| Sample with variation in depression symptoms<br>N = 27,297 |  |  |  |  | Sample with variation in anxiety symptoms<br>N = 26,452 |  |  |  |
| --- | --- | --- | --- | --- | --- | --- | --- | --- |
| Variable | Overall Percent |  | Between Percent | Within Percent | Overall Percent |  | Between Percent | Within Percent |
| Financial adversity index |  |  |  |  |  |  |  |  |
| 0 | 90.85% |  | 98.16% | 90.33% | 90.63% |  | 98.11% | 90.13% |
| 1 | 6.40% |  | 33.72% | 22.68% | 6.54% |  | 34.35% | 22.72% |
| 2 | 2.09% |  | 13.91% | 19.34% | 2.14% |  | 14.21% | 19.36% |
| 3 | 0.58% |  | 4.12% | 20.67% | 0.60% |  | 4.22% | 20.63% |
| 4-5 | 0.09% |  | 0.74% | 19.89% | 0.09% |  | 0.76% | 19.82% |
|  | Overall Mean | Overall SD | Between SD | Within SD | Overall Mean | Overall SD | Between SD | Within SD |
| Depression symptoms | 5.52 | 5.65 | 5.20 | 2.73 | 5.64 | 5.69 | 5.20 | 2.77 |
| Anxiety symptoms | 4.18 | 4.98 | 4.59 | 2.42 | 4.31 | 5.01 | 4.58 | 2.46 |

### Associations between financial adversity and mental health

Increases in financial adversity were negatively associated with depression (Figure 1a) and anxiety (Figure 1b) symptoms. These associations were dose-dependent (Supplemental Table S4); the largest increases in both outcomes were seen for participants experiencing 4-5 financial adversities (incidence rate ratio [IRR] = 1.36; 95% confidence interval [CI] = 1.31 to 1.41] for depression symptoms; IRR = 1.42; 95% CI = 1.37 to 1.48 for anxiety), whilst people experiencing one financial adversity had smaller increased risk compared to people with zero financial adversities (IRR = 1.12; 95% CI = 1.12 to 1.13 for depression; IRR = 1.15; 95% CI = 1.15 to 1.16 for anxiety).

**Figure 1.**
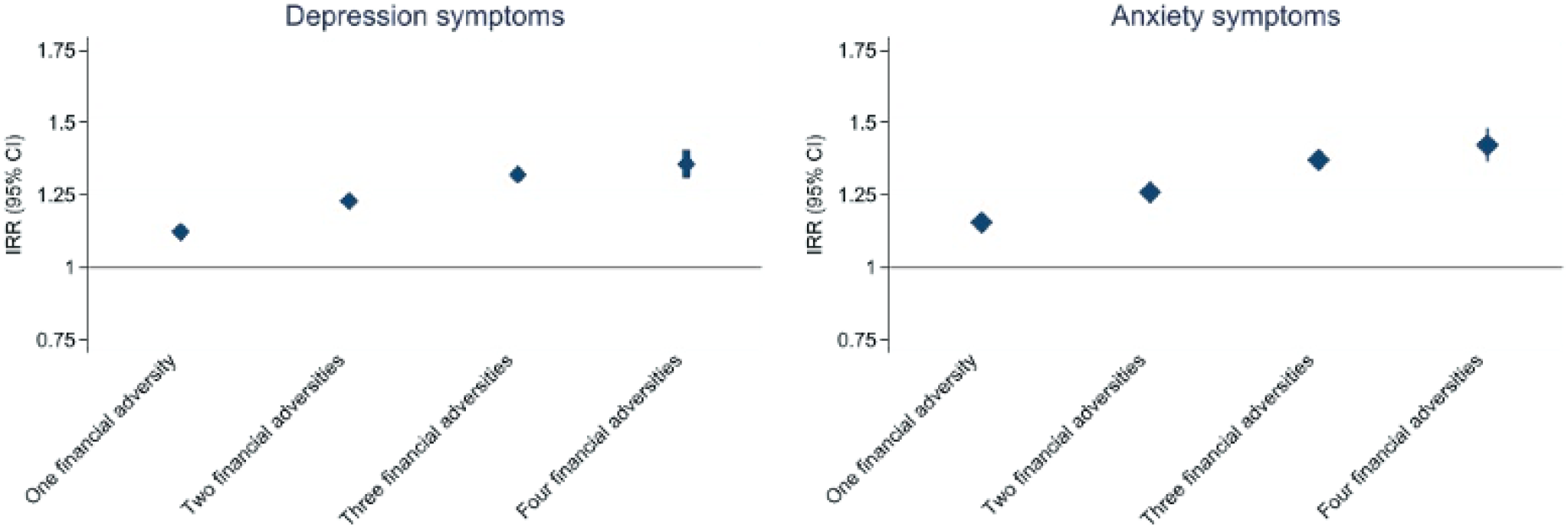

### Interactions with the use of financial supports

Accessing financial supports had little or no impact on mental health (Table 3). Specifically, having used any type of financial support at least once over the study period had a small positive impact on mental health when participants had experienced only one financial adversity. However, this interaction was in the other direction in the cases of two to three financial adversities. The direct impact of financial adversities on mental health remained when interactions with financial supports were included in the model (see Supplemental Table S4 for direct effects).

**Table 3.** Interactions between the financial adversity index and the use of financial supports with depression and anxiety symptoms, derived from fixed effects Poisson regression models, weighted

|  | Depression symptoms<br>N = 27,297 |  |  | Anxiety symptoms<br>N = 26,452 |  |  |
| --- | --- | --- | --- | --- | --- | --- |
|  | IRR | 95% CI |  | IRR | 95% CI |  |
| Interaction between any financial support and... |  |  |  |  |  |  |
| One financial adversity | **0.98** | **0.97** | **0.99** | **0.98** | **0.96** | **0.99** |
| Two financial adversities | **1.03** | **1.01** | **1.06** | **1.02** | **1.00** | **1.05** |
| Three financial adversities | **1.08** | **1.02** | **1.14** | 1.02 | 0.96 | 1.09 |
| Four financial adversities | 0.92 | 0.70 | 1.22 | 1.23 | 0.86 | 1.77 |
| Interaction between any charity based support and... |  |  |  |  |  |  |
| One financial adversity | **0.92** | **0.91** | **0.94** | **0.92** | **0.90** | **0.94** |
| Two financial adversities | **0.91** | **0.88** | **0.94** | **0.90** | **0.87** | **0.94** |
| Three financial adversities | **0.91** | **0.87** | **0.95** | **0.87** | **0.82** | **0.91** |
| Four financial adversities | **0.86** | **0.79** | **0.94** | **0.85** | **0.77** | **0.94** |
| Interaction between any government based support and... |  |  |  |  |  |  |
| One financial adversity | **0.98** | **0.97** | **0.99** | 0.99 | 0.98 | 1.00 |
| Two financial adversities | 1.01 | 0.99 | 1.03 | 1.01 | 0.99 | 1.03 |
| Three financial adversities | 1.00 | 0.97 | 1.03 | 1.01 | 0.97 | 1.05 |
| Four financial adversities | 1.00 | 0.91 | 1.10 | 1.07 | 0.96 | 1.18 |
| Interaction between any work based support and... |  |  |  |  |  |  |
| One financial adversity | **1.03** | **1.02** | **1.04** | **1.03** | **1.02** | **1.04** |
| Two financial adversities | **1.02** | **1.00** | **1.04** | **1.03** | **1.01** | **1.06** |
| Three financial adversities | **1.05** | **1.02** | **1.09** | **1.06** | **1.02** | **1.10** |
| Four financial adversities | 0.98 | 0.91 | 1.06 | 0.98 | 0.90 | 1.07 |
| Interaction between any other support and.... |  |  |  |  |  |  |
| One financial adversity | **0.94** | **0.93** | **0.95** | **0.95** | **0.93** | **0.96** |
| Two financial adversities | 1.01 | 0.99 | 1.04 | 1.01 | 0.98 | 1.03 |
| Three financial adversities | 0.98 | 0.95 | 1.02 | **0.94** | **0.91** | **0.98** |
| Four financial adversities | 0.97 | 0.90 | 1.04 | 0.94 | 0.87 | 1.02 |

Note. Data were weighted to the proportions of gender, age, ethnicity, country, and education obtained from the Office for National Statistics. Models adjusted for time-varying covariates. Statistics for direct effects terms are presented in Supplemental Table S6.

Charity based support (the use of food banks or clothing donations) was the only specific form of financial support to consistently attenuate the impact of financial adversity on depression and anxiety symptoms (Table 3). There was also evidence that the magnitude of this attenuation increased with a greater number of financial adversities.

Government based support (e.g., Universal Credit) had a very small impact on depression symptoms, but only in the presence of one financial adversity. Work based support (sick pay or furlough) exacerbated the impact of financial adversity on depression and anxiety symptoms but had no impact on people who had experienced four to five financial adversities. The use of other financial supports (e.g., bank loans or insurance claims) only attenuated the impact of financial adversity on depression and anxiety symptoms when one financial adversity was present (and not more).

### Sensitivity analyses

When sick pay was removed from the work based financial support variable, furlough exacerbated the impact of financial adversity on mental health but had no impact when people had experienced four to five financial adversities (Supplemental Table S5).

## Discussion

Increasing exposure to financial adversity had a strong dose-dependent negative relationship with mental health over time during the COVID-19 pandemic, even when accounting for time-varying adversities such as infection with COVID-19 and not being able to access essentials and loneliness. This echoes findings from other population based studies conducted during the COVID-19 pandemic.^6,12^ There was little evidence that financial supports attenuated this adverse relationship: although having used at least one of any of the financial supports measured had a small impact on mental health in people with only one financial adversity, it actually exacerbated the impact in people who had experienced more than one such adversity. Work based support (sick pay and in particular furlough), also exacerbated the impact. This is congruent with other research conducted in the early stages of the pandemic which found that furlough associated with declines in mental wellbeing.^4^

Charity based supports (foodbanks and clothing donations) were the only financial support to attenuate the impact of financial adversity on mental health in people with increasing numbers of adversities. Foodbank use, often used by the severely food insecure, is associated with poorer self-rated health, mental health disorders and disability prior to the pandemic.^13^ It is possible that in our study, charity based support attenuated the impact of financial adversity on mental health by supplementing nutritional needs that would otherwise not have been met and that are linked to mental health via physical health. This could have been due to pandemic-related food shortages or extreme financial hardship, although our analyses controlled for not being able to access sufficient food and in people who had experienced four to five adversities, charity based supports no longer benefited mental health.

This study has many strengths, including a large heterogenous sample, repeated monthly measurements over two years, and robust statistical methods which used sampling weights. However, although we included assessments of financial support use at four different time points over a two year period, we did not examine timing of their use, which may have been important for mental health given the last minute nature of the announcement of the Coronavirus Job Retention Scheme extension for example. Second, although large and heterogenous, and made more reflective of the general UK population by the use of sampling weights, probability sampling methods were not used, and the study is therefore not representative. We were also unable to examine associations between pre-pandemic financial adversity, financial supports, and mental health due to a lack of data. Findings may therefore have been affected by factors related to the COVID-19 pandemic and not generalisable to other circumstances.

Our findings imply that most forms of financial support are inadequate for alleviating the impact of financial adversity on mental health. This is cause for concern given the expected upcoming recession;^14^ the impact on mental health may be substantial. Overall, findings suggest that more comprehensive forms of support for people experiencing financial adversity are needed to reduce the impact on mental health, particularly in people experiencing multiple financial adversities.

## Data Availability

The dataset analysed for the current study, as well as study documentation and codebook are publicly available from the UK Data Service (UCL COVID-19 Social Study, 2020-2022). Statistical code is available upon request from Elise Paul.

https://beta.ukdataservice.ac.uk/datacatalogue/studies/study?id=9001

## Statements

### Data availability statement

The UCL COVID-19 Social Study documentation and codebook are available for download at https://osf.io/jm8ra/. Statistical code is available upon request from Elise Paul.

### Ethics approval

The study was approved by the University College London Research Ethics Committee (approval number 12467/005).

## Acknowledgements

The researchers are grateful for the support of a number of organisations with their recruitment efforts including: the UKRI Mental Health Networks, Find Out Now, UCL BioResource, SEO Works, FieldworkHub, and Optimal Workshop.

## Supplemental Materials

### Recruitment methods

The sample was recruited using three primary approaches. First, convenience sampling was used, including promoting the study through existing networks and mailing lists (including large databases of adults who had previously consented to be involved in health research across the UK), print and digital media coverage, and social media. Second, more targeted recruitment was undertaken focusing on (i) individuals from a low-income background, (ii) individuals with no or few educational qualifications, and (iii) individuals who were unemployed. Third, the study was promoted via partnerships with third sector organisations (e.g., charities or community sector organisations) to include marginalised or vulnerable groups including adults with pre-existing mental health conditions, older adults, carers, and people experiencing domestic violence or abuse.

#### Outcome variables

For both scales, respondents rated the frequency of symptoms over the past week on a scale from 0 (not at all) to 3 (nearly every day). Symptoms were summed (depression symptoms range: 0-27; anxiety symptoms range: 0-21). When study data collection was weekly (1 April to 31 August 2020), the maximum depression and anxiety scores per month were used, instead of averages, in order to maintain consistency with depression and anxiety scores later in the study, when data were collected monthly, and to ease interpretation of results (i.e., averages would result numbers with decimals).

#### Financial adversity

Due to the low number of participants who had experienced all five financial adversities in a given week, scores were recorded as 0, 1, 2, 3, or 4+. Because the work based support variable included both sick pay and furlough, we conducted analyses with the furlough variable only to avoid including people who were ill and therefore more likely to be experiencing depression and anxiety symptoms.

#### Time-varying covariates1

Loneliness was measured using a short form of the Revised UCLA Loneliness Scale (UCLA-R) and also included as a time-varying covariate. Each item was rated from 0 (hardly ever) to 2 (often), with higher scores indicating more loneliness in the past week. When data collection was weekly, the maximum loneliness score per month was used.

### Statistical Analysis

#### Fixed effects regression

In the fixed effects approach, individuals serve as their own reference point, which accounts for any confounding associations between time-invariant (stable) covariates such as genetic predisposition, personality traits, and history of mental health difficulties between predictors and outcomes.^9^

The basic fixed effects regression model can be expressed as follows:

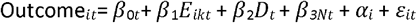

where Outcome_*it*_ is a measure of individual *i*’s self-harm thoughts or self-harm behaviours at time *t*, E is individual *i*’s predictor variable at time *t, D*_*t*_ is a vector of indicator variables for day, *N*_*t*_ is a continuous variable for days since lockdown, is unobserved time invariant confounding factors, and *ε* is error.

**Table S1.** Financial support stem questions and dates of administration

| Stem question | Date of administration |
| --- | --- |
| Have you used any of the following since lockdown began? | 28 May 2020 |
| Have you used any of the following in the last two months? | 30 July 2020 |
| Have you used any of the following SINCE THE START OF AUGUST? | 23 December 2020 |
| Have you used any of the following SINCE THE START OF 2021? | 9 September 2021 |

**Table S2.** Comparison of included and excluded participants, unweighted

|  |  | Excluded from depression symptoms sample<br>N = 40,135 | Included in depression symptoms sample<br>N = 27,297 | Inclusion |  |  | Excluded from anxiety symptoms sample<br>N = 40,980 | Included in anxiety symptoms sample<br>N = 26,252 | Inclusion |  |  |
| --- | --- | --- | --- | --- | --- | --- | --- | --- | --- | --- | --- |
|  |  |  |  | OR | 95% CI |  |  |  | OR | 95% CI |  |
| Gender | Male | 27.81% | 25.11% | Ref |  |  | 28.80% | 24.38% | Ref |  |  |
|  | Female | 72.19% | 74.89% | **1.14** | **1.13** | **1.15** | 71.20% | 75.62% | **1.21** | **1.19** | **1.22** |
| Age | 60+ | 42.41% | 39.31% | Ref |  |  | 43.96% | 38.22% | Ref |  |  |
|  | 46-59 | 32.09% | 35.37% | **1.22** | **1.20** | **1.24** | 31.44% | 35.90% | **1.30** | **1.28** | **1.32** |
|  | 30-45 | 19.98% | 20.72% | **1.14** | **1.12** | **1.16** | 19.35% | 21.15% | **1.21** | **1.19** | **1.23** |
|  | 18-29 | 5.52% | 4.60% | **0.94** | **0.92** | **0.96** | 5.25% | 4.74% | **1.01** | **0.98** | **1.03** |
| Ethnicity | White | 95.67% | 96.55% | Ref |  |  | 95.76% | 96.53% | Ref |  |  |
|  | Ethnic minority groups | 4.33% | 3.45% | **0.79** | **0.77** | **0.81** | 4.24% | 3.47% | **0.78** | **0.76** | **0.80** |
| Education | Degree or higher | 67.51% | 70.77% | Ref |  |  | 67.12% | 71.13% | Ref |  |  |
|  | A-levels or equivalent | 17.26% | 16.54% | **0.93** | **0.92** | **0.94** | 17.36% | 16.45% | **0.91** | **0.90** | **0.92** |
|  | Up to GCSE | 15.23% | 12.69% | **0.82** | **0.81** | **0.83** | 15.52% | 12.42% | **0.79** | **0.78** | **0.81** |
| Household income | ≥ £30k | 57.41% | 59.77% | Ref |  |  | 57.06% | 60.07% | Ref |  |  |
|  | < £30k | 42.59% | 40.23% | 0.99 | 0.98 | 1.00 | 42.94% | 39.93% | **0.97** | **0.96** | **0.98** |
| Long term physical health condition (ref none) |  | 44.13% | 42.12% | **0.98** | **0.97** | **0.99** | 44.30% | 41.95% | **0.98** | **0.97** | **0.99** |
| Long term mental health condition (ref none) |  | 16.86% | 16.25% | **1.04** | **1.03** | **1.06** | 16.11% | 16.71% | **1.04** | **1.03** | **1.06** |
| Financial adversity index | 0 | 88.42% | 90.66% | Ref |  |  | 88.87% | 90.45% | Ref |  |  |
|  | 1 | 7.88% | 6.53% | **0.84** | **0.83** | **0.86** | 7.58% | 6.67% | **0.85** | **0.84** | **0.87** |
|  | 2 | 2.70% | 2.15% | **0.83** | **0.80** | **0.85** | 2.58% | 2.20% | **0.83** | **0.80** | **0.86** |
|  | 3 | 0.84% | 0.58% | **0.74** | **0.70** | **0.79** | 0.80% | 0.59% | **0.74** | **0.69** | **0.78** |
|  | 4-5 | 0.17% | 0.09% | **0.57** | **0.49** | **0.66** | 0.17% | 0.09% | **0.56** | **0.48** | **0.65** |
| Unable to access essentials |  | 3.57% | 2.51% | **0.82** | **0.80** | **0.85** | 3.42% | 2.58% | **0.80** | **0.78** | **0.83** |
| Death or serious illness in other |  | 7.12% | 6.51% | **0.96** | **0.94** | **0.98** | 6.89% | 6.64% | 0.99 | 0.97 | 1.01 |
| Physical or psychological abuse |  | 6.71% | 5.94% | **0.99** | **0.96** | **1.01** | 6.37% | 6.13% | 0.98 | 0.96 | 1.01 |
| Infection with COVID-19 |  | 4.45% | 2.61% | **0.59** | **0.58** | **0.61** | 4.31% | 2.64% | **0.59** | **0.58** | **0.61** |
| Loneliness |  | 4.86 | 4.83 | **1.03** | **1.02** | **1.03** | 4.79 | 4.87 | **1.04** | **1.04** | **1.05** |
| Depression symptoms |  | 6.05 | 5.53 | **0.98** | **0.98** | **0.98** | 5.77 | 5.69 | **0.99** | **0.99** | **0.99** |
| Anxiety symptoms |  | 4.53 | 4.16 | **1.00** | **1.00** | **1.00** | 4.30 | 4.30 | **1.00** | **1.00** | **1.01** |

Note. Bold indicates $p < .05$ . Results are from two logistic regression models which mutually adjusted for all variables and with inclusion in the depression and anxiety symptoms samples as the outcome variables, respectively.

**Table S3.**
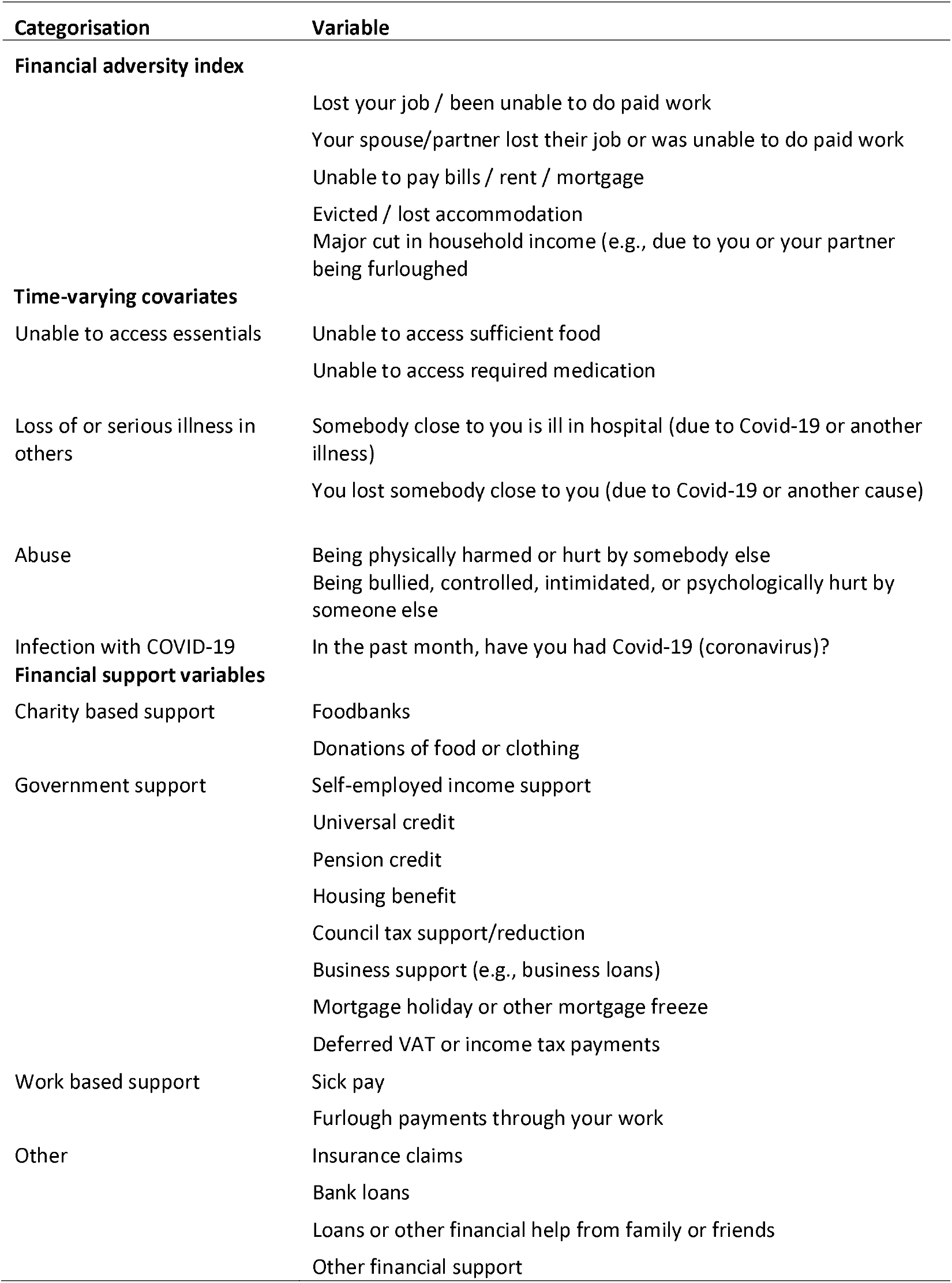
Categorisation of study developed items

**Table S4.**
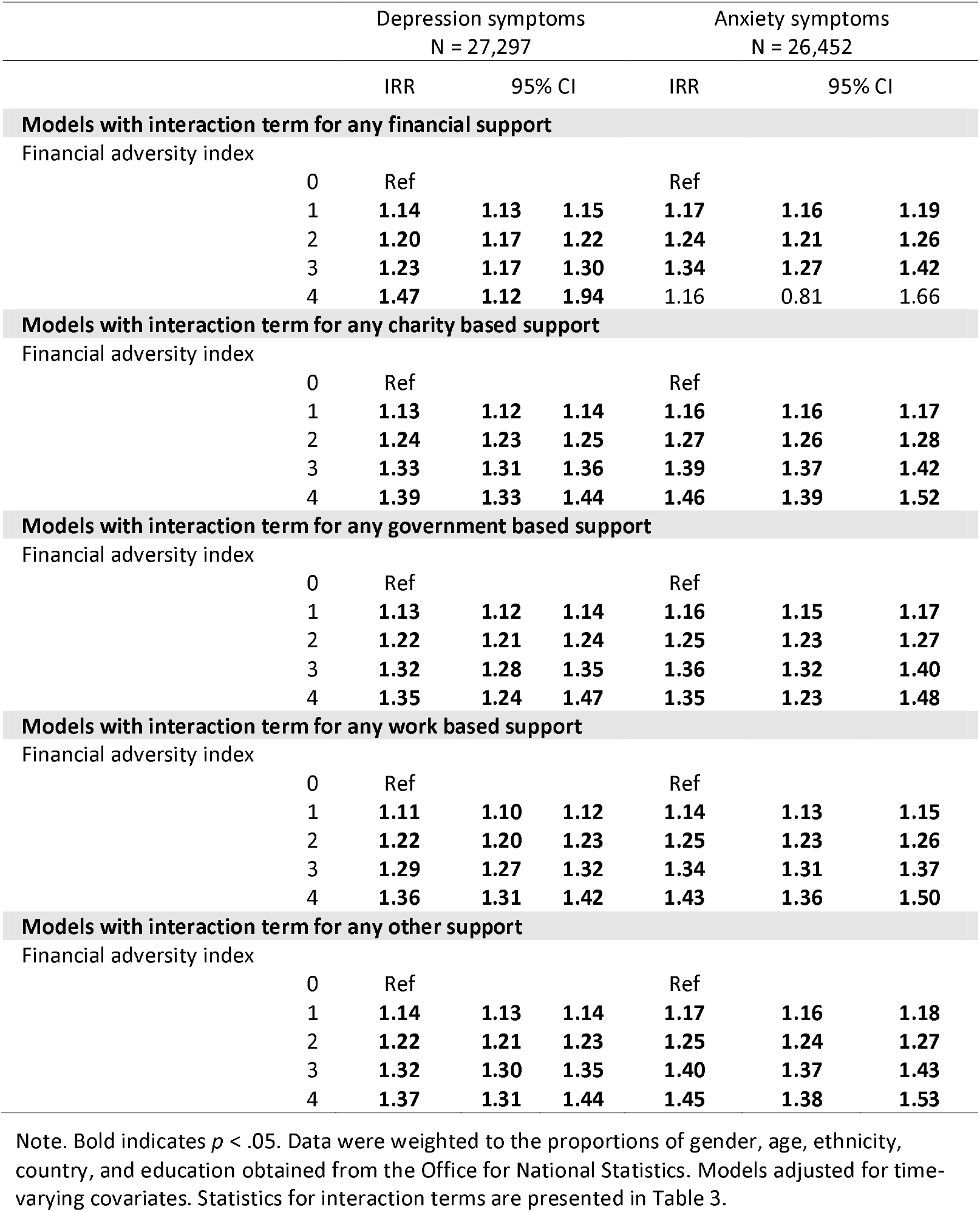
Direct associations between financial adversity and depression and anxiety symptoms from fixed effects Poisson regression models which included interaction terms, weighted

|  |  | Depression symptoms<br>N = 27,297 |  |  | Anxiety symptoms<br>N = 26,452 |  |  |
| --- | --- | --- | --- | --- | --- | --- | --- |
|  |  | IRR | 95% CI |  | IRR | 95% CI |  |
| Models with interaction term for any financial support |  |  |  |  |  |  |  |
| Financial adversity index |  |  |  |  |  |  |  |
|  | 0 | Ref |  |  | Ref |  |  |
|  | 1 | 1.14 | 1.13 | 1.15 | 1.17 | 1.16 | 1.19 |
|  | 2 | 1.20 | 1.17 | 1.22 | 1.24 | 1.21 | 1.26 |
|  | 3 | 1.23 | 1.17 | 1.30 | 1.34 | 1.27 | 1.42 |
|  | 4 | 1.47 | 1.12 | 1.94 | 1.16 | 0.81 | 1.66 |
| Models with interaction term for any charity based support |  |  |  |  |  |  |  |
| Financial adversity index |  |  |  |  |  |  |  |
|  | 0 | Ref |  |  | Ref |  |  |
|  | 1 | 1.13 | 1.12 | 1.14 | 1.16 | 1.16 | 1.17 |
|  | 2 | 1.24 | 1.23 | 1.25 | 1.27 | 1.26 | 1.28 |
|  | 3 | 1.33 | 1.31 | 1.36 | 1.39 | 1.37 | 1.42 |
|  | 4 | 1.39 | 1.33 | 1.44 | 1.46 | 1.39 | 1.52 |
| Models with interaction term for any government based support |  |  |  |  |  |  |  |
| Financial adversity index |  |  |  |  |  |  |  |
|  | 0 | Ref |  |  | Ref |  |  |
|  | 1 | 1.13 | 1.12 | 1.14 | 1.16 | 1.15 | 1.17 |
|  | 2 | 1.22 | 1.21 | 1.24 | 1.25 | 1.23 | 1.27 |
|  | 3 | 1.32 | 1.28 | 1.35 | 1.36 | 1.32 | 1.40 |
|  | 4 | 1.35 | 1.24 | 1.47 | 1.35 | 1.23 | 1.48 |
| Models with interaction term for any work based support |  |  |  |  |  |  |  |
| Financial adversity index |  |  |  |  |  |  |  |
|  | 0 | Ref |  |  | Ref |  |  |
|  | 1 | 1.11 | 1.10 | 1.12 | 1.14 | 1.13 | 1.15 |
|  | 2 | 1.22 | 1.20 | 1.23 | 1.25 | 1.23 | 1.26 |
|  | 3 | 1.29 | 1.27 | 1.32 | 1.34 | 1.31 | 1.37 |
|  | 4 | 1.36 | 1.31 | 1.42 | 1.43 | 1.36 | 1.50 |
| Models with interaction term for any other support |  |  |  |  |  |  |  |
| Financial adversity index |  |  |  |  |  |  |  |
|  | 0 | Ref |  |  | Ref |  |  |
|  | 1 | 1.14 | 1.13 | 1.14 | 1.17 | 1.16 | 1.18 |
|  | 2 | 1.22 | 1.21 | 1.23 | 1.25 | 1.24 | 1.27 |
|  | 3 | 1.32 | 1.30 | 1.35 | 1.40 | 1.37 | 1.43 |
|  | 4 | 1.37 | 1.31 | 1.44 | 1.45 | 1.38 | 1.53 |

Note. Bold indicates $p < .05$ . Data were weighted to the proportions of gender, age, ethnicity, country, and education obtained from the Office for National Statistics. Models adjusted for time-varying covariates. Statistics for interaction terms are presented in Table 3.

**Table S5.** Sensitivity analysis: interactions between financial adversity and furlough with depression and anxiety symptoms from fixed effects Poisson regression models, weighted

|  |  | Depression symptoms<br>N = 27,297 |  |  | Anxiety symptoms<br>N = 26,452 |  |  |
| --- | --- | --- | --- | --- | --- | --- | --- |
|  |  | IRR | 95% CI |  | IRR | 95% CI |  |
| Financial adversity index |  |  |  |  |  |  |  |
|  | 0 | Ref |  |  | Ref |  |  |
|  | 1 | **1.11** | **1.10** | **1.12** | **1.14** | **1.13** | **1.15** |
|  | 2 | **1.21** | **1.20** | **1.22** | **1.24** | **1.23** | **1.26** |
|  | 3 | **1.29** | **1.27** | **1.32** | **1.35** | **1.32** | **1.38** |
|  | 4-5 | **1.35** | **1.30** | **1.41** | **1.43** | **1.37** | **1.50** |
| Interaction between furlough use and... |  |  |  |  |  |  |  |
|  | One financial adversity | **1.05** | **1.03** | **1.06** | **1.04** | **1.03** | **1.06** |
|  | Two financial adversities | **1.06** | **1.04** | **1.09** | **1.05** | **1.03** | **1.08** |
|  | Three financial adversities | **1.07** | **1.03** | **1.11** | **1.06** | **1.02** | **1.10** |
|  | Four-five financial adversities | 1.00 | 0.92 | 1.09 | 0.97 | 0.88 | 1.06 |

Note. Bold indicates $p < .05$ . Data were weighted to the proportions of gender, age, ethnicity, country, and education obtained from the Office for National Statistics. Models adjusted for time-varying covariates.

## References

1. Kiely KM, Leach LS, Olesen SC, Butterworth P. How financial hardship is associated with the onset of mental health problems over time. Soc Psychiatry Psychiatr Epidemiol. 2015 Jun 1;50(6):909–18.

2. Guerin RJ, Barile JP, Thompson WW, McKnight-Eily L, Okun AH. Investigating the Impact of Job Loss and Decreased Work Hours on Physical and Mental Health Outcomes Among US Adults During the COVID-19 Pandemic. J Occup Environ Med. 2021 Sep;63(9):e571.

3. NatCen Social Research. Exploring the relationship between economic security, furlough and mental distress [Internet]. 2021 p. 5. Available from: https://dera.ioe.ac.uk/38381/1/Exploring-the-relationship-between-economic-security-furlough-and-mental-distress.pdf

4. Wels J, Booth C, Wielgoszewska B, Green MJ, Di Gessa G, Huggins CF, et al. Mental and social wellbeing and the UK coronavirus job retention scheme: Evidence from nine longitudinal studies. Soc Sci Med. 2022 Sep 1;308:115226.

5. Donnelly R, Farina MP. How do state policies shape experiences of household income shocks and mental health during the COVID-19 pandemic? Soc Sci Med. 2021 Jan 1;269:113557.

6. Wickham S, Bentley L, Rose T, Whitehead M, Taylor-Robinson D, Barr B. Effects on mental health of a UK welfare reform, Universal Credit: a longitudinal controlled study. Lancet Public Health. 2020;5(3):e157–64.

7. Löwe B, Kroenke K, Herzog W, Gräfe K. Measuring depression outcome with a brief self-report instrument: sensitivity to change of the Patient Health Questionnaire (PHQ-9). J Affect Disord. 2004;81(1):61–6.

8. Spitzer RL, Kroenke K, Williams JB, Löwe B. A brief measure for assessing generalized anxiety disorder: the GAD-7. Arch Intern Med. 2006;166(10):1092–7.

9. Office for National Statistics. Population estimates for the UK, England and Wales, Scotland and Northern Ireland [Internet]. 2020 May [cited 2020 Sep 30]. Available from: https://www.ons.gov.uk/peoplepopulationandcommunity/populationandmigration/populationestimates/bulletins/annualmidyearpopulationestimates/mid2018

10. Hainmueller J, Xu Y. Ebalance: A Stata package for entropy balancing. J Stat Softw. 2013;54(7).

11. Stata Corp. Stata Statistical Software: Release 17. College Station, TX: StataCorp LLC; 2021.

12. Donnelly R, Zajdel R, Farina MP. Inequality in Household Job Insecurity and Mental Health: Changes During the COVID-19 Pandemic. Work Occup. 2022 Sep 13;07308884221123255.

13. Loh S, Knight A, Loopstra R. Working-age adults using food banks in England have significantly poorer health and higher rates of mental health conditions than adults in the general population: A cross-sectional quantitative study. Health Soc Care Community. 2021;29(5):1594–605.

14. HM Treasury. Forecasts for the UK economy: October 2022 [Internet]. 2022. Available from: https://www.gov.uk/government/statistics/forecasts-for-the-uk-economy-october-2022

## References

1. Russell D. UCLA Loneliness Scale (Version 3): Reliability, validity, and factor structure. J Pers Assess [Internet]. 1996 [cited 2020 Oct 29];66(1). Available from: https://www.tandfonline.com/doi/abs/10.1207/s15327752jpa6601_2

